# Strengthening response coordination through public health emergency operations centers in Africa: Lessons learned from 56-week webinar sessions, 2020-2021

**DOI:** 10.1101/2022.11.22.22282627

**Authors:** Womi Eteng, Abrham Lilay, Senait Tekeste, Wessam Mankoula, Emily Collard, Chimwemwe Waya, Emily Rosenfeld, Chuck Menchion Wilton, Martin Muita, Liz McGinley, Yan Kawe, Ali Abdullah, Ariane Halm, Jian Li, Virgil L Lokossou, Youssouf Kanoute, Ibrahima Sonko, Merawi Aragaw

## Abstract

**Background:** Following the declaration of coronavirus disease 2019 (COVID-19) as a pandemic on 11 March 2020, in-person events including trainings were canceled to limit the spread of the pandemic. A virtual learning program was established in May 2020 by Africa Centers for Disease Control and Prevention, the World Health Organization, and other partners to strengthen COVID-19 response coordination through the public health emergency operations centers (PHEOCs). We present a review of the webinar series, the experience, and the lessons learned.

**Method:** A data extraction tool was developed to retrieve data from the Africa CDC’s webinar data repository. Major findings were synthesized and described per thematic area.

**Results:** A total of 12,715 (13% of the 95,230 registrants) attended the 56 PHEOC webinar sessions between June 2020 and December 2021 and 47% of the attendees came from 17 countries. Of those who attended, 8,528 (70%) were from Africa. The webinars provided 97 learning hours with an average length of 1.18 hours per session. On average, there were 235 attendees per session. In addition, there was an average of 26 interactions between participants and facilitators per session. A total of 4,084 (44%) of the participants (9,283) responded to the post-session surveys, with over 95% rating the webinar topics as being relevant to their work, contributed to improving their understanding of PHEOC operationalization, and with extensive ease of comprehension.

**Conclusion:** The virtual training served the intended audience given the high number of participants from African member states, with satisfactory feedback on training relevance. We highlighted a just-in-time, progressively adaptive experience in delivering a PHEOC/PHEM virtual learning in Africa with a consequential global audience at the peak of the COVID-19 pandemic.

## Introduction

On March 11, 2020, the World Health Organization (WHO) declared coronavirus disease 2019 (COVID-19) a pandemic[1]. The COVID-19 pandemic severely tested existing multi-sectoral coordination structures and mechanisms for health emergency information and resource management by the public health emergency operations centers (PHEOCs) or similar institutions in countries across the African continent and globally. The pandemic further revealed the need for a whole-of-government approach including stakeholders within and outside the traditional health sector arrangements. The timely implementation of functional PHEOCs has been documented to be a factor for improved response to emergencies[2, 3]. A functional PHEOC is equally important in meeting the minimum requirements of the International Health Regulations (IHR-2005)[4]. The PHEOC employs an Incident Management System (IMS), which is a standardized, scalable, and flexible emergency management structure with sets of procedures and protocols to provide a coordinated approach while avoiding duplication of effort for all types of health emergencies.

With the COVID-19 pandemic, programmatic plans and events including in-person training were disrupted due to the travel and physical distancing measures implemented globally to limit the spread of the pandemic[5]. Virtual systems and communication became the alternative business norm for organizing meetings and delivery of trainings and other activities. This option became the leverage for delivering webinars accessible through smartphones, tablets, and computers from any part of the world. Preferred technology had platforms that support audio and video capabilities[6]. A webinar allows various interactive opportunities, such as discussion, instant messaging functions, conducting polls, surveys, and knowledge checks. In addition, an interactive and accessible online community of practice can be established alongside webinar training sessions, offering a space to share presentations, continued discussion, future learning, and country-to-country experiential learning and mentoring. Training, learning, and satisfaction through online webinars could lead to excellent outcomes compared to face-to-face, as concluded in a meta-analysis conducted by Gegenfurtner et al.[7].

A virtual learning platform was established in May 2020 to build and/or strengthen capacity in PHEOC management for a coordinated COVID-19 response by creating an online Community of Practice (CoP) comprising mainly PHEOC professionals from the African continent. The platform was designed to address the immediate knowledge gaps, facilitate the exchange of experience and serve as a springboard for launching a sustained post-COVID-19 experiential learning base. The primary target audience was personnel working in PHEOCs across the African continent. However, participants across the globe were actively engaged throughout the weekly webinars. A complementary Discord™ Platform was established to provide continuous engagement and networking opportunities[8].

To lead the overall process of the weekly PHEOC Webinar Series, a core working group was established with members identified from Africa Centers for Disease Control and Prevention (Africa CDC), World Health Organization Regional Office for Africa (WHO AFRO), World Health Organization Regional Office for the Eastern Mediterranean (WHO EMRO), WHO Headquarters, Public Health England (PHE), US Centers for Disease Control and Prevention (US CDC), European Centers for Disease Prevention and Control (ECDC), Resolve to Save Lives (RTSL) and Emergency Operations Centre Network (EOC-NET).

A secretariat was established at the Africa CDC Headquarters to oversee program coordination, including scheduling and coordination of meetings, producing technical briefs, and developing publicity materials (flyers and broadcast emails). The secretariat also provided an enquiry desk to address queries and respond to requests for information or technical assistance. The country experience was often included in the webinars where guest presenters from National PHEOCs were invited to share their experience of PHEOC management and operations during the pandemic era.

The approach to selecting webinar topics evolved, hence remained dynamic and adaptive. In the initial stages, the first few webinar topics were identified based on the results of a very quickly administered baseline survey that was distributed to those in the PHEOC field that were known to the PHEOC partners working group. With continuous interactions with webinar participants, through direct feedback, survey findings, and consultations with key actors at the country level, the webinar topics were tailored toward the needs and interests of participants. As the webinars progressed, participants’ inquiries into previously treated topics prompted the re-organization of one-off topics into series to provide a deep-dive learning experience into specific thematic areas of public health emergency management (PHEM) including PHEOC implementation. Participants who completed the module series and knowledge checks were rewarded with e-certificates of completion. Presenters were typically senior public health specialists, PHEOC professionals, and program leads from across the continent and beyond who covered topics based on their areas of expertise.

A review of the webinar series was conducted to document the experience acquired and identify recommendations that could potentially serve as a future reference for the establishment of virtual learning programs and CoP within the continent of Africa and in settings similar to Africa.

## Methods

### Data sources

The relevant documents for the 56-Webinar Series held on Thursdays from 3:00 p.m. East Africa Time (EAT) were retrieved from the shared drive webinar data repository created by Africa CDC, WHO, and other partners. These documents consisted of presentations, post-webinar polls and survey findings, and webinar performance reports.

### Relevant variables and Data analysis

Key indicators were chosen to assess webinar performance before, during, and after the event. Relevant registration and participation information were extracted and consolidated for analysis using a pre-defined data extraction too. These included attendance rates from Africa and around the world per webinar, the countries with the most attendees, the length of the webinar sessions, and the number of participants-to-facilitator discussions.

Participants’ responses to the post-webinar polls, which began during webinar 16 and administered until webinar 52, were reviewed, with an additional question including clarifying the attendee’s role in the PHEOC from webinar 30 to 52. The questions were:

- Will you attend another webinar based on the delivery of this session?
- Did the session improve your understanding of the topic?
- Was the content easy to follow?
- Was there enough time for questions and discussion?
- Would you recommend the session to someone?
- How relevant was the topic to your current role in the COVID-19 response?
- Is your current role related to Public Health Emergency Operations Centre?

A thematic analysis (i.e., inductive, and semantic approaches) was used to group, analyze, and describe major findings. Further descriptive statistics (percentages, tables, charts, etc.) were used to summarize the quantitative data.

### Ethics Statement

This paper is a review, not research involving patients, and it does not require institutional review approval. The data is anonymized, and no personal information is included.

## Results

### Webinar participation

A total of 95,230 participants were registered and 12,715 (13%) attended the PHEOC Webinar Series held between June 2020 and December 2021. Of those who attended, 8,528 (67%) were from the African continent. Over the webinar period, an average of 33 with a range of 8 to 50 countries participated from the African continent (**Fig 1**).

**Fig. 1.**
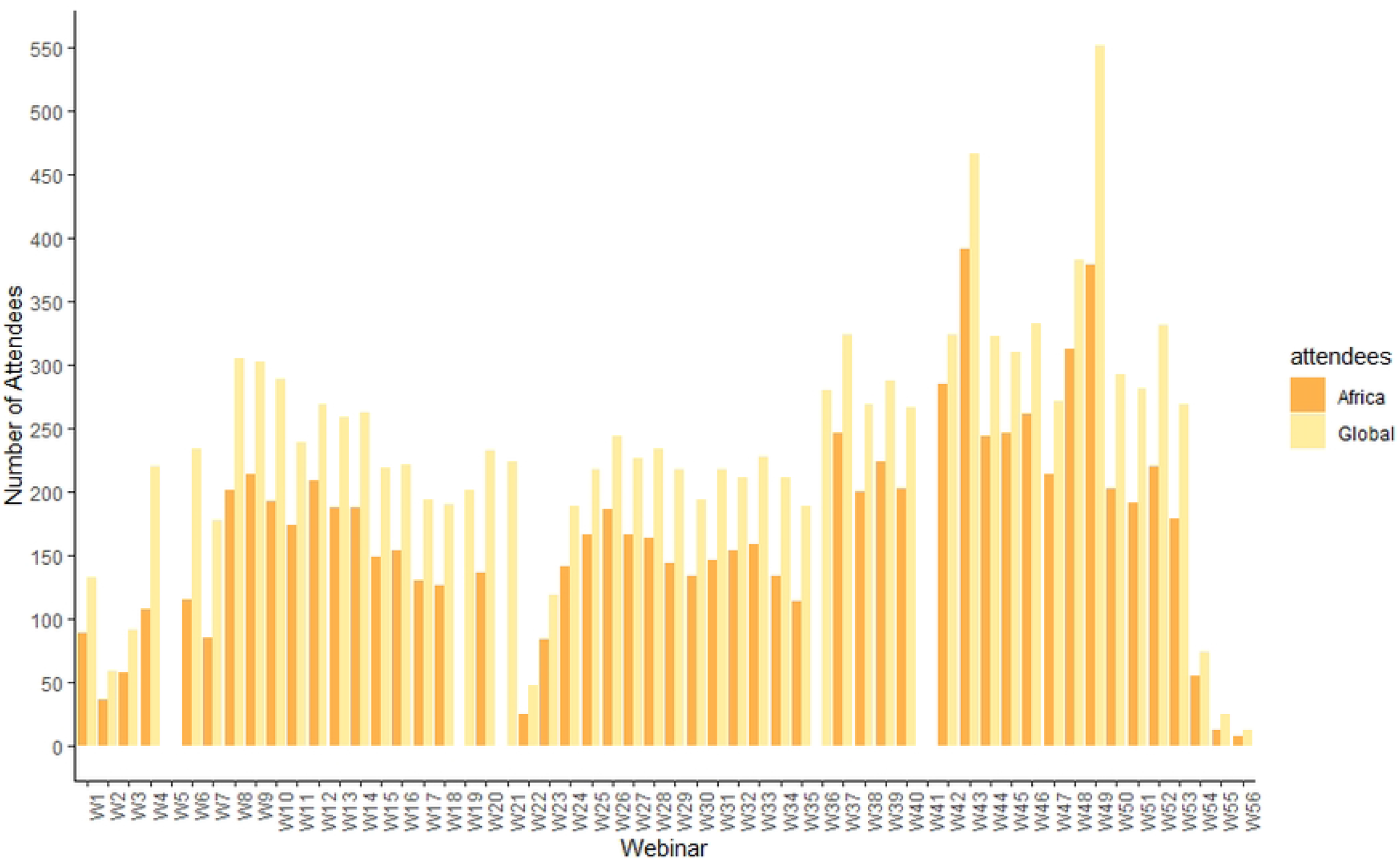

Seventeen countries were identified throughout the webinar periods as having the top three attendance rankings based on the number of attendees each week, with 13 being from the African continent. Nigeria, the United States of America (USA), Kenya, and Ethiopia accounted for 45% of all webinar attendees (**Fig 2**).

**Fig. 2.**
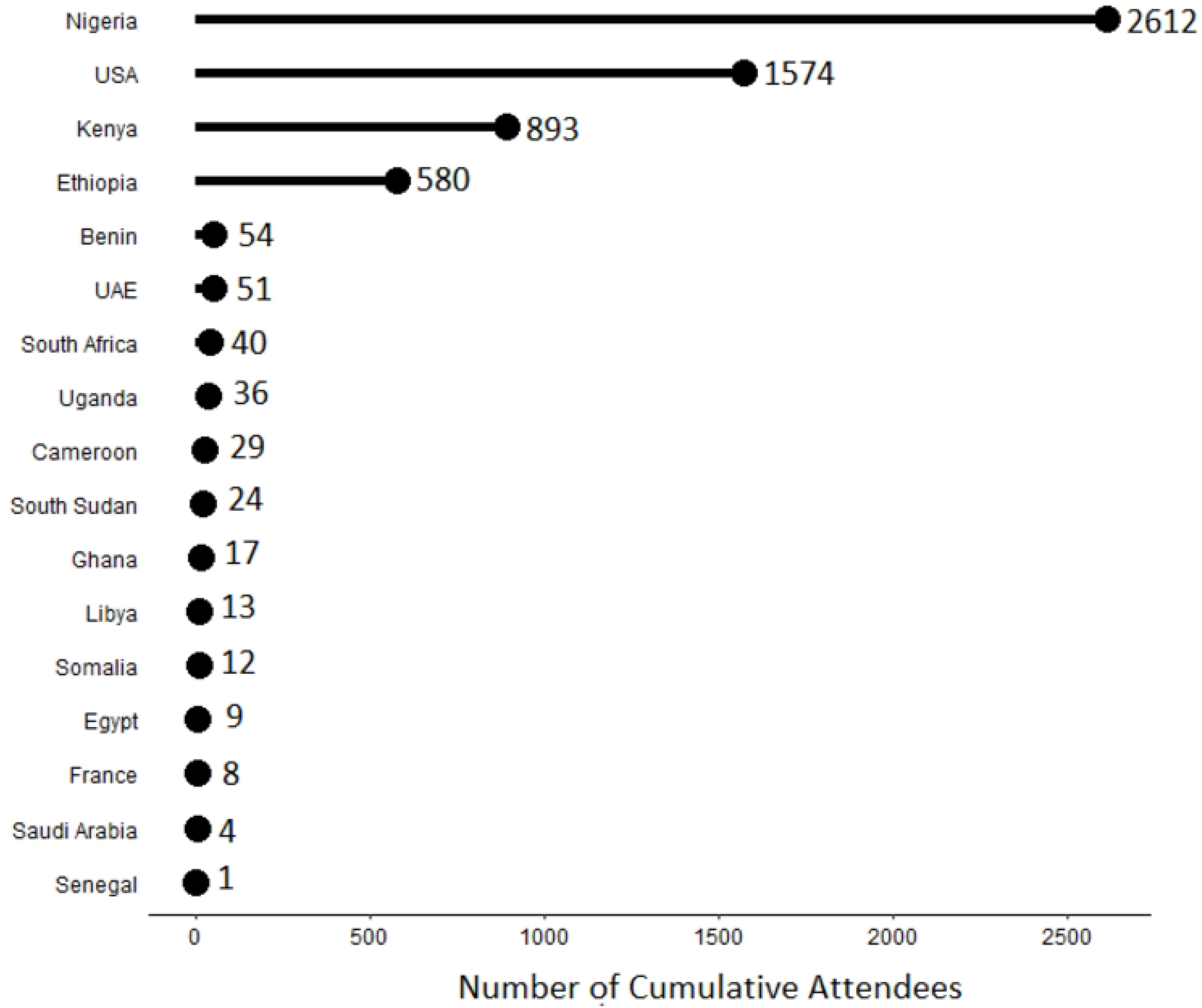

### Establishing the Webinars and Community of Practice

#### Baseline survey

In May 2020 before the first webinar, an initial baseline survey was created using an online survey tool and shared with stakeholders from across the PHEOC networks. The responses to the survey were used to help develop and refine the topic and content of the webinars.

The summary of survey responses showed the working group what challenges Member States were facing and also areas of good practice in PHEOCs responding to COVID-19 (**Table 1**).

**Table 1:**
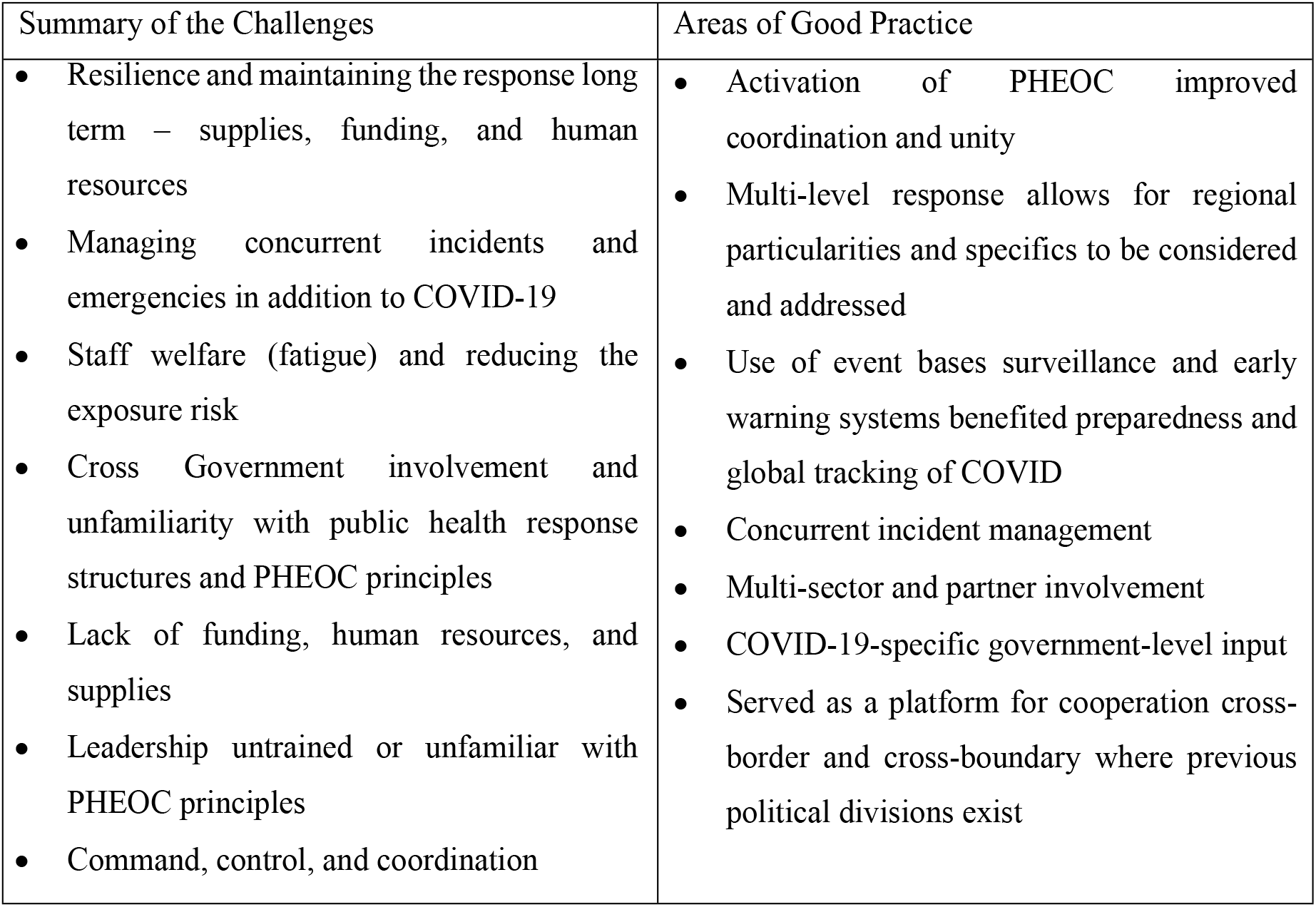
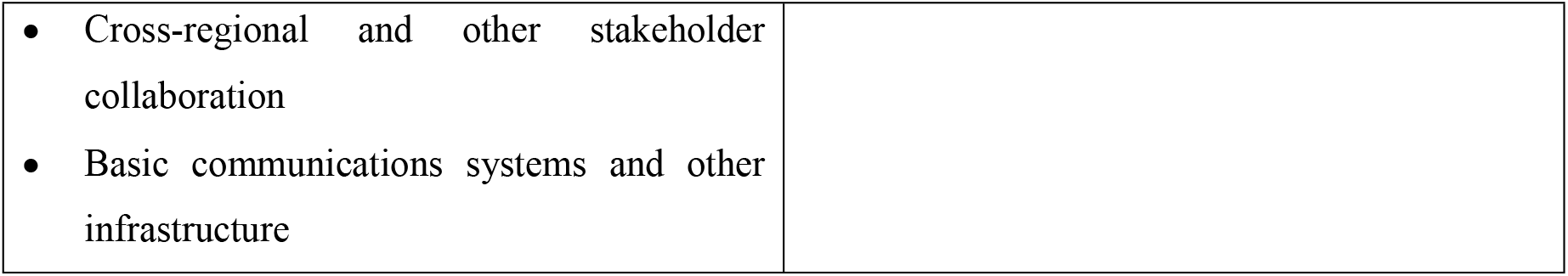
Summary response of baseline survey, May 2020

| Summary of the Challenges | Areas of Good Practice |
| --- | --- |
| • Resilience and maintaining the response long term – supplies, funding, and human resources<br>• Managing concurrent incidents and emergencies in addition to COVID-19<br>• Staff welfare (fatigue) and reducing the exposure risk<br>• Cross Government involvement and unfamiliarity with public health response structures and PHEOC principles<br>• Lack of funding, human resources, and supplies<br>• Leadership untrained or unfamiliar with PHEOC principles<br>• Command, control, and coordination | • Activation of PHEOC improved coordination and unity<br>• Multi-level response allows for regional particularities and specifics to be considered and addressed<br>• Use of event bases surveillance and early warning systems benefited preparedness and global tracking of COVID<br>• Concurrent incident management<br>• Multi-sector and partner involvement<br>• COVID-19-specific government-level input<br>• Served as a platform for cooperation cross-border and cross-boundary where previous political divisions exist |

|  |
| --- |
| • Cross-regional and other stakeholder collaboration<br>• Basic communications systems and other infrastructure |

#### Webinar Planning and Coordination

A secretariat was established at the Africa CDC Headquarters to oversee program coordination, including scheduling and coordination of working group meetings, producing technical briefs, and developing publicity materials (flyers and broadcast emails).

The PHEOC Partners working group had the responsibility for developing and agreeing on the weekly webinar topic, schedule, and content. Decisions were generally made by consensus, and where not possible the chair of the group (Africa CDC or WHO) would make the final decision. Through direct feedback, survey findings, and consultations with key actors at the country level, the webinar topics were tailored toward the needs and interests of participants. The secretariat was then responsible for the official marketing of the webinars with the development of a flyer which was shared through agreed PHEOC networks, email distribution lists, and word of mouth.

#### Quality assurance of webinar content

The PHEOC Partners working group was responsible for ensuring consistency of presentations with established terminologies, workflow, and alignment with existing guidance documents, except where innovative approaches are considered. Presenters were required to share final presentations along with any related reference documents prior to the webinar for review and agreement by the working group.

#### Webinar delivery

The Zoom© platform was chosen to host the weekly webinar series which was administered by Africa CDC as part of their secretariat role. Each webinar was moderated and supported by a small team of approximately 4 people. The lead moderator was responsible for leading the participants through the webinar, introducing the speakers, and facilitating group discussions and Q&A sessions. The other members of the team supported the lead moderator by monitoring the instant messages, providing expert input for Q&A sessions, signposting to relevant resources, and backing up the moderator in case needed. In addition to this Africa, CDC had IT support on hand in case of any technical issues and the latter webinars were supported by translators for French & Arabic.

#### Webinar topics and series

During the early phases, topics for the webinars were identified and prioritized by the core team using a majority vote system to identify and prioritize topics. However, survey findings and consultations with key public health actors at the country level were used to inform webinar topics tailored to the interests of participants in later months. The prioritized topics were arranged into a series to allow participants to take a deep dive into the content and gain a thorough understanding of each topic.

During the webinar period, 56 topics were covered across eight thematic areas. Each of the Incident Management System (IMS) and incident leadership series had 14 topics. The information management series covered seven topics, while the multi-sectoral coordination, PHEOC handbook, and legal framework series each covered five topics. Other series had four or fewer topics (**Fig 3**).

**Fig. 3.**
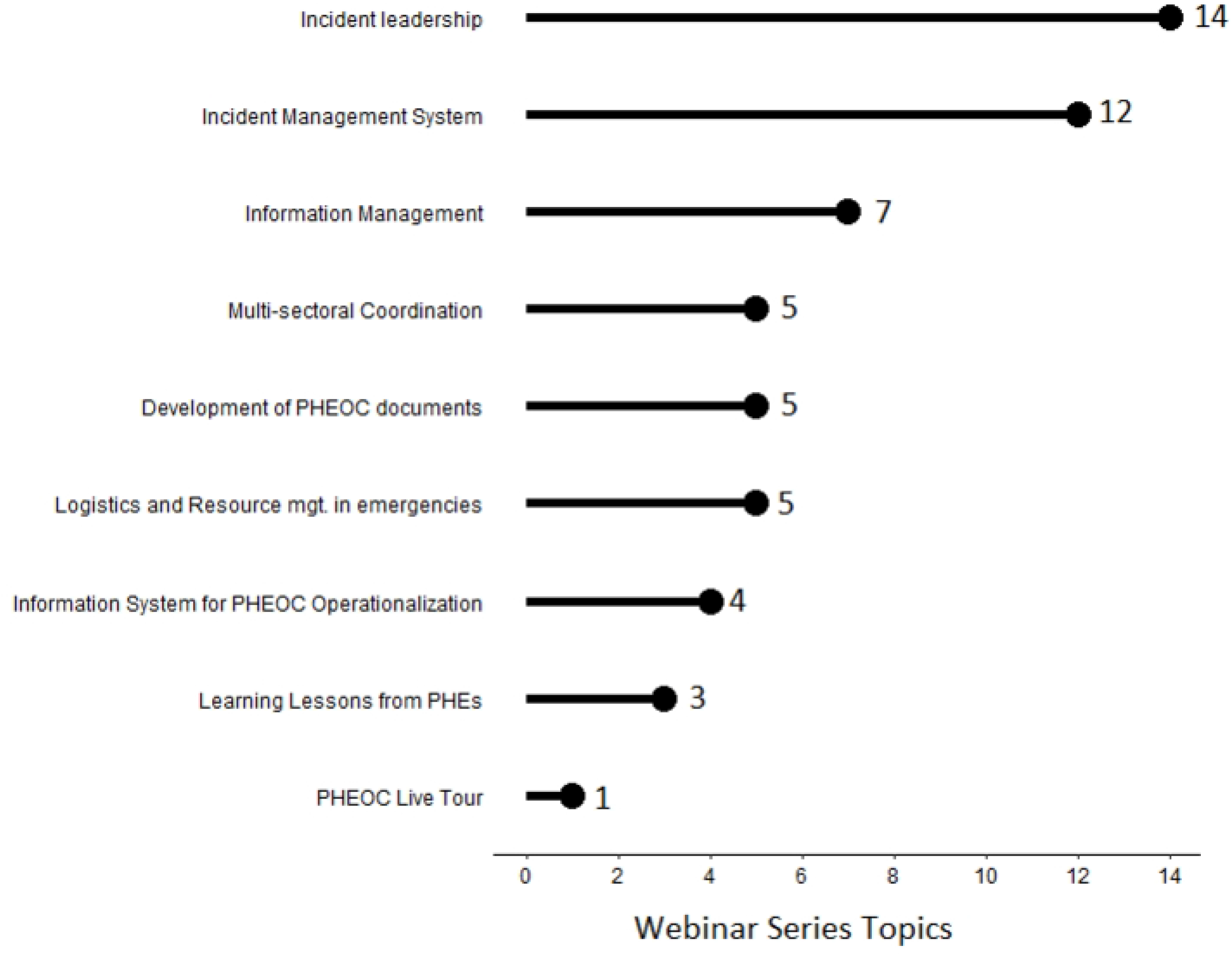

Development of PHEOC document part 2, IMS round-up, and development of PHEOC documents part 1 sessions were attended by 551 (4.3%), 466 (3.7%), and 382 (3.0%) participants, respectively. On the other side, 47 (0.4%), 25 (0.2%), and 12 (0.1%) attended Watch Mode operations: EBS tools, PHEOC information systems part 3, and PHEOC information systems part 4 (**Fig 4**).

**Fig. 4.**
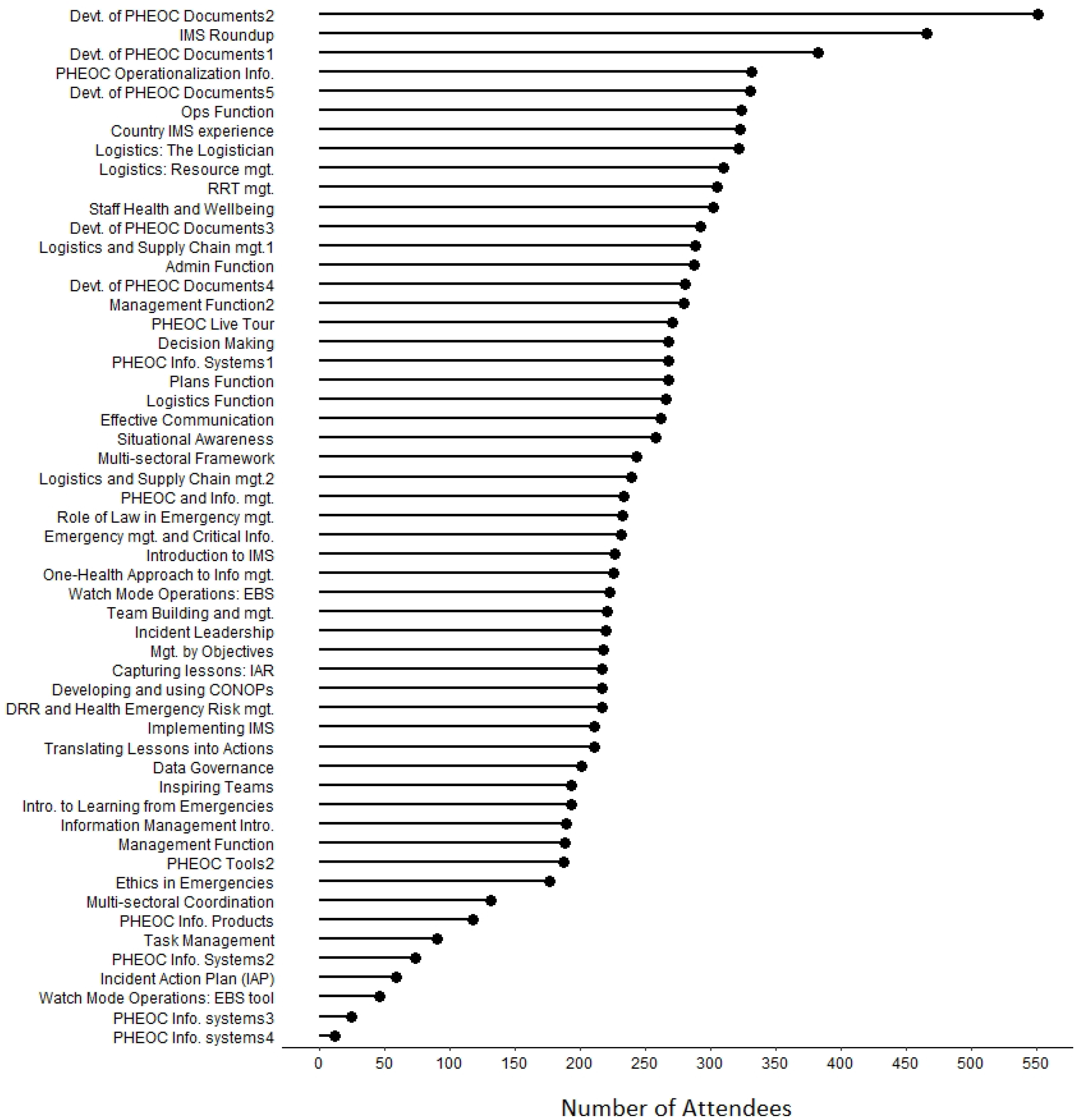

#### Post-webinar survey

During the period when the post-webinar surveys were administered, 4,084 (44%) of the webinar participants (9,283) responded. Over 95% responded positively to the topic’s relevance to their current role, the likelihood of recommending a topic to another colleague, the session’s contribution to improving their understanding of the issue, interest in attending another session, and content that is easy to understand. Furthermore, 85% said there was enough time for discussion at the end of the sessions. From webinars 30 to 52, where the additional question was added, 2,404 (81%) of the 2970 webinar participants responded that they had a role related to PHEOC (**Table 2**).

**Table 2:**
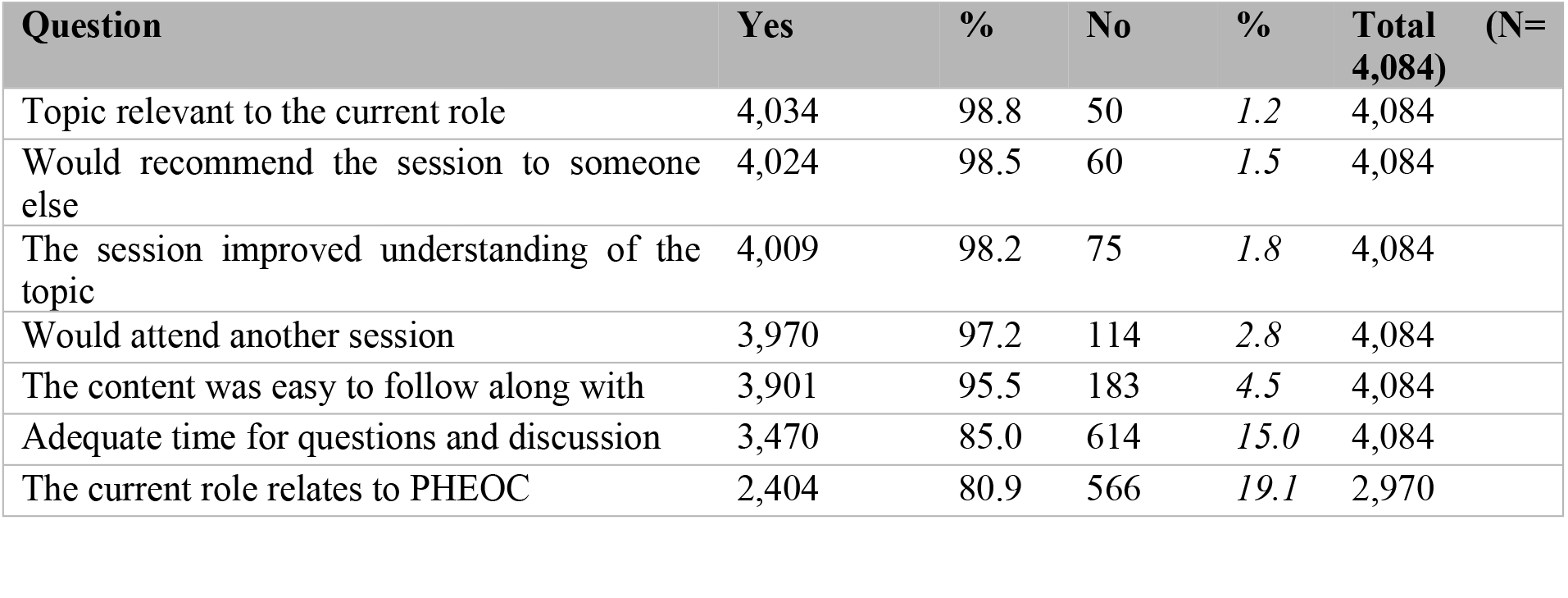
Responses of post-webinar surveys, 2020-2021

| Question | Yes | % | No | % | Total (N= 4,084) |
| --- | --- | --- | --- | --- | --- |
| Topic relevant to the current role | 4,034 | 98.8 | 50 | 1.2 | 4,084 |
| Would recommend the session to someone else | 4,024 | 98.5 | 60 | 1.5 | 4,084 |
| The session improved understanding of the topic | 4,009 | 98.2 | 75 | 1.8 | 4,084 |
| Would attend another session | 3,970 | 97.2 | 114 | 2.8 | 4,084 |
| The content was easy to follow along with | 3,901 | 95.5 | 183 | 4.5 | 4,084 |
| Adequate time for questions and discussion | 3,470 | 85.0 | 614 | 15.0 | 4,084 |
| The current role relates to PHEOC | 2,404 | 80.9 | 566 | 19.1 | 2,970 |

#### Interaction and networking within the online CoP

There were 1,407 interactions between webinar participants and facilitators, including Q&A and opinion sharing throughout the webinar period. In the individual webinar sessions, there were, on average, 26 interactions between the participants and facilitators, ranging from 4 to 56 interactions per session.

The ‘Discord Platform’ was acknowledged by members of the online CoP to be an effective platform that enabled continuous engagement and networking across Member States, and increased their knowledge and skills on the topics addressed. It facilitated peer-to-peer learning through country-to-country, PHEOC-to-PHEOC communication about implementing coordinated COVID-19 responses. The platform was utilized by participants, before, during, and after webinars sessions, to discuss real-time issues and needs (e.g., advocacy for decision-makers commitment, PHEOC funding, and legal framework development, etc.), sharing of experience, lessons learned and best practices (e.g., PHEOC-to-PHEOC mentorship on potential PHEOCs support in COVID-19 response), and required documentations for PHEOC operationalization (e.g., sharing of relevant PHEOC manuals, SOPs, training materials and events, and other useful references).

As of September 2022, about 1789 online community members were found active on the platform. The core working group members continuously work to improve and ensure that the online CoP and the virtual training through Webinar Series endure beyond the COVID-19 pandemic.

### Reward on Participation

To encourage participation and intake of training as well as to reward active participants, e-certificates were provided to those who attended all the webinar sessions and completed the knowledge check/quizzes. A total of 2,102 participants (15% of those who attended the certificated series) received e-certificates, with logos of all coordinating partners.

## Discussion

The study presented findings from the review of a virtual networking and learning platform providing bi/weekly capacity building webinar series, from 2020-2021. The establishment of such a platform was driven by the aim of providing just-in-time training in PHEOC operationalization as hubs supporting the implementation of coordinated COVID-19 responses in countries across Africa.

The outcomes of the review showed the virtual networking and learning platform, and related CoP outreached the intended primary audience. The Webinar series significantly contributed to building and/or strengthening the knowledge of PHEOCs professionals through active networking, and country-to-country sharing of experience, lessons learned and best practices, and required documentation for implementing PHEOCs and coordinated COVID-19 responses. However, these findings should be interpreted with caution due to the limitations of the review and Webinar series.

There were sufficient registrants (over 95,000) interested in joining the PHEOC Webinar Series. However, there was an overall low attendance rate (13%), and close to half (47%) of the total webinar participants came from only 17 countries. This high registration rate may reflect the genuine need and interest in PHEOC management material at the time, conversely, the low attendance rate may reflect several scenarios including a lack of time to attend a 1–2-hour long webinar during the COVID-19 pandemic, particularly for those working in a PHEOC, a lack of technological infrastructure to join the webinar online; conflicting commitments at the time of the webinars.

Most (67% of 12,715) of the attendees across all webinars were from the African continent (Benin, Cameroon, Egypt, Ethiopia, Ghana, Kenya, Libya, Nigeria, Senegal, Somalia, South Africa, South Sudan, and Uganda).

Nigeria, the United States of America (USA), Kenya, and Ethiopia accounted for 45% of all webinar attendees. This strong representation from the African continent and the United States could be interpreted on several levels including that there was a greater need for information and learning on PHEOC management and operationalization from these. However, the inclusion of key partners of Africa CDC and US CDC in the PHEOC Webinar Series working group is likely to have resulted in a biased in advertisement/word of mouth in engagement in the webinars, resulting in higher numbers of participants from these in particular.

The adaptive and evolving nature of the webinar program helped to improve the delivery of the sessions and tailored it to the interests of participants across the continent and beyond. The average number of attendees (235) per session was good and about 70% of the webinar sessions had at least 200 attendees. This could be because the webinar topics were tailored to their interests and the fact that there were increasingly strong planning and coordination efforts between the working group and attendees/community of practice as the webinars continued.

Inviting countries to share their experiences in the management of COVID-19 helped other countries learn and adapt the lessons in their context and to engage in an active exchange with other experts in the region. Through the process, 17 countries from Africa and outside the continent had a chance to share their experiences. The PHEOC webinar sessions containing the country experience sharing were mainly provided with the help of PowerPoint presentations by Subject Matter Experts from invited countries and various public health organizations. It found that resource persons (74%) give lectures during the online program with help of PowerPoint presentations[9].

Based on the findings from this review, the average webinar length was 1.8 hours, with sessions lasting anywhere from 1.5 to 2.7 hours, this reflected the complexity of the topic discussed and the engagement of the attendees in the subsequent Q&A session. Organizers recognized that participants tended to drop off the webinar after one hour, likely because of conflicting commitments, therefore, it is recommended that future webinar sessions do not extend beyond one hour. And where it is likely to last longer up to 1.5 hours, it may be beneficial to inform participants ahead of time and keep them actively engaged[10]. The PHEOC webinar sessions were held on Thursdays, taking the recommendations of a study that attributed better participation in attending webinars on Tuesdays, Wednesdays, and Thursdays, these being the best days for live events in countries where Saturday and Sunday are the weekends[11]. In the first 12 months of the series, sessions were conducted at the same time each Thursday, on reflection and based on feedback from participants, it was decided that weekly sessions should use an alternating time to mitigate the chance of a clash with re-occurring pre-existing meetings.

Almost all respondents (over 95%) to the post-webinar surveys agreed that the topics of the webinar sessions were relevant, the contents were easy to comprehend and contributed to improving their understanding. The working group met regularly (virtually) and was in frequent contact via email to discuss and refine the weekly content, using expertise from across all partner organizations. Likewise, the working group was open to suggestions from international colleagues and attendees regarding on-topic content and presentation of content. An active and efficient secretariat was crucial in the development and distribution of Webinar brochures and links which were communicated via registrants’ email and published on the websites of the Africa CDC. Previous studies have indicated that where respondents (89%) revealed that when organizers provide webinar links well in advance for registration and expressed (91%) that organizers have conducted webinars on relevant topics[9]. In addition, another study found that webinar participants (75%) were interested in the topic rather than the speaker or the company organizing the webinar[11]. The success of the topic choice and strong communication of webinar sessions in advance is indicative of the commitment of the PHEOC Network partners, PHEOC Webinar working group, and secretariat, and emphasizes the effort involved in conducting a weekly, interactive webinar series, especially when hosted by SME’s whom themselves were engaged in COVID-19 response activities. The amount of effort involved in organizing similar public health learning tools/models should be taken into account by planning teams, and consideration given to the resources available, the ambitiousness of the scale of the learning program, and the level of commitment required.

In terms of the time taken per session, only 85% of attendees agreed that enough time was allotted at the end of the webinar session for discussion and answering questions. There were only 1,407 interactions between webinar participants and organizers during the webinar period, including Q&A and opinion sharing, resulting in an average of 26 interactions per session. The interactions observed appear to be low as compared to the number of attendees per session (235 on average), which could be due to the limited time allocated for discussion, however other considerations may include the likelihood of attendees listening to the session while conducting other work, language barriers or their ability or confidence to engage with the other attendees and facilitators in a virtual environment.

## Limitation

As a major limitation, we considered the low post-webinar survey response rate (below half of the participants), missing data on participants’ relevant experiences, and might have biased the topic selection process in favor of the numerical strength of respondents who, though working within PHEOCs, are not occupying critical roles to immediately translate the learning into practice.

## Conclusions

In conclusion, the learning program fulfilled its purpose of addressing knowledge gaps, especially on PHEOC operationalization and its role in the effective coordination of responses to PHEs, providing training opportunities, and facilitating exchanges during the pandemic, which was well received by the target audience in the continent and also benefited professionals across the globe. We found that there was a good number of participants per webinar session and participants were satisfied with the content and relevance of the webinar topics. On the other side, the results revealed that there were some concerns with the time allocated for discussions at the end of the session. Besides, there was an overall low attendance rate though the high number of registrants indicates interest and acceptance of the Webinar Series, and almost half of the participants came from just a few countries, possibly explained by biased in the advertisement of the webinars based on partners involved. Hence, we recommend continuing the learning platform in the African continent and similar settings to Africa. We believe that some modifications to address the gaps and to put a robust virtual learning platform in place should be considered during the planning process including time allocation for discussion and strengthening communication with relevant stakeholders at regional and country levels to ensure a high participation rate.

## Data Availability

Data could be accessed upon request to the author

## Contributors

WE and AL conducted relevant article searches, developed a data extraction tool, analyzed the data, and wrote the draft manuscript. WE, AL, ST, WM, EC, CW, ER, CM, MM, LM, YK, AA, AH, JL, VL, YK, IS, and MA contributed to the writing and reviewed the final manuscript. All authors read and approved the final manuscript.

## Availability of data and materials

The data used in the article are available and the author will provide access upon request.

## Acknowledgement

We would like to acknowledge all countries that attended the webinar sessions and participants for completing the feedback survey and for continued support throughout the webinar period.

